# Investigating phenotypes of pulmonary COVID-19 recovery – a longitudinal observational prospective multicenter trial

**DOI:** 10.1101/2021.06.22.21259316

**Authors:** Thomas Sonnweber, Piotr Tymoszuk, Sabina Sahanic, Anna Boehm, Alex Pizzini, Anna Luger, Christoph Schwabl, Manfred Nairz, Katharina Kurz, Sabine Koppelstätter, Magdalena Aichner, Bernhard Puchner, Alexander Egger, Gregor Hoermann, Ewald Wöll, Günter Weiss, Gerlig Widmann, Ivan Tancevski, Judith Löffler-Ragg

**Affiliations:** Department of Internal Medicine II, Medical University of Innsbruck, Innsbruck, Austria; Department of Radiology, Medical University of Innsbruck, Innsbruck, Austria; The Karl Landsteiner Institute, Reha Zentrum Münster, Münster, Austria; Central Institute of Medical and Chemical Laboratory Diagnostics, University Hospital Innsbruck, Innsbruck, Austria; MLL Munich Leukemia Laboratory, Munich, Germany; Department of Internal Medicine, St. Vinzenz Hospital, Zams, Austria; Data Analytics As a Service Tirol, daas.tirol, Innsbruck, Austria

**Keywords:** computed tomography, cluster analysis, COVID-19, dyspnea, long COVID, lung damage, machine learning, post-COVID-19 syndrome, recovery phenotype, respiratory sequelae, SARS-CoV-2

## Abstract

**Background:** COVID-19 is associated with long-term pulmonary symptoms and may result in chronic pulmonary impairment. The optimal procedures to prevent, identify, monitor, and treat these pulmonary sequelae are elusive.

**Research question:** To characterize the kinetics of pulmonary recovery, risk factors and constellations of clinical features linked to persisting radiological lung findings after COVID-19.

**Study design and methods:** A longitudinal, prospective, multicenter, observational cohort study including COVID-19 patients (n = 108). Longitudinal pulmonary imaging and functional readouts, symptom prevalence, clinical and laboratory parameters were collected during acute COVID-19 and at 60-, 100- and 180-days follow-up visits. Recovery kinetics and risk factors were investigated by logistic regression. Classification of clinical features and study participants was accomplished by k-means clustering, the k-nearest neighbors (kNN), and naive Bayes algorithms.

**Results:** At the six-month follow-up, 51.9% of participants reported persistent symptoms with physical performance impairment (27.8%) and dyspnea (24.1%) being the most frequent. Structural lung abnormalities were still present in 45.4% of the collective, ranging from 12% in the outpatients to 78% in the subjects treated at the ICU during acute infection. The strongest risk factors of persisting lung findings were elevated interleukin-6 (IL6) and C-reactive protein (CRP) during recovery and hospitalization during acute COVID-19. Clustering analysis revealed association of the lung lesions with increased anti-S1/S2 antibody, IL6, CRP, and D-dimer levels at the early follow-up suggesting non-resolving inflammation as a mechanism of the perturbed recovery.

Finally, we demonstrate the robustness of risk class assignment and prediction of individual risk of delayed lung recovery employing clustering and machine learning algorithms.

**Interpretation:** Severity of acute infection, and systemic inflammation is strongly linked to persistent post-COVID-19 lung abnormality. Automated screening of multi-parameter health record data may assist the identification of patients at risk of delayed pulmonary recovery and optimize COVID-19 follow-up management.

**Clinical Trial Registration:** ClinicalTrials.gov: NCT04416100

## Introduction

The ongoing COVID-19 pandemic challenges health care systems worldwide. As of June 2021, the John Hopkins dashboard^1^ reports 178 million global cases and 3.8 million COVID-19-related deaths^2^. Although the vast majority of COVID-19 patients display mild disease, approximately 10-15% of cases progress to a severe condition and approximately 5% suffer from critical illness^3,4^. Similar to severe acute respiratory syndrome (SARS), a significant portion of COVID-19 patients report lingering or recurring clinical impairment and cardiopulmonary recovery may take several months to years^5–11^. This observation has led to the introduction of the term ‘long COVID’, defined by persistence of COVID-19 symptoms for more than four weeks, and the ‘post-COVID-19 syndrome’ referring to symptom persistence for more than twelve weeks^12,13^. Evidence-based strategies for prediction, monitoring and treatment of post-acute COVID-19 sequelae are urgently needed. We herein prospectively analyzed prevalence of non-resolving lung abnormalities, risk factors and clinical feature sets associated with delayed pulmonary recovery during the first six months of COVID-19 convalescence and tested whether a multi-parameter machine learning approach may help discerning subjects at risk of persistent lung damage.

## Methods

### Study design

The CovILD (“Development of interstitial lung disease in COVID-19”) study^5^ was initiated in April 2020. Adult residents of Tyrol, Austria, with typical clinical presentation and a positive SARS-CoV-2 PCR test from a nasal or oropharyngeal swab^14^ were enrolled by three centers: Department of Internal Medicine II at the Medical University of Innsbruck (primary follow-up center), St. Vinzenz Hospital in Zams and the acute rehabilitation facility in Münster. During the 2020 SARS-CoV2 outbreaks, the regional health system was able to guarantee the best standard of care including intensive therapy and mechanical ventilation if necessary. None of the participants received corticosteroids as a therapy of the acute infection.

In total 190 COVID-19 patients were screened for study participation. N = 18 subjects denied to give an informed consent, N = 27 were declared difficulties to appear at the study follow-ups. Of the 145 enrolled participants, 37 were excluded from analysis due to an incomplete data record precluding classification analyses (Supplementary Figure S1).

All participants gave written informed consent. The study was approved by the institutional review board at the Medical University of Innsbruck (approval number: 1103/2020), and registered at ClinicalTrials.gov (NCT04416100).

### Procedures

We retrospectively assessed patient characteristics during acute COVID-19 and performed follow-up investigations at 60 days (63 ± 23 days (mean ± SD); visit 1), 100 days (103 ± 21); visit 2) and 180 days (190 ± 15; visit 3) after the diagnosis of COVID-19. Each visit included clinical examination, assessment of symptoms and performance status with a standardized questionnaire, lung function testing, capillary blood gas analysis, trans-thoracic echocardiography, and low-dose computed tomography (CT) scan of the chest. CT scans were evaluated for the presence of ground-glass opacities (GGO), consolidations, bronchial dilation, and reticulations as defined by the Fleischner society. Lung findings were graded with a CT severity score (0-25 points), as previously published^5^.

Lung function impairment was defined by at least one of the following: (1) forced vital capacity (FVC) < 80% predicted, (2) forced expiratory volume in 1 second (FEV_1_) < 80% predicted, FEV1:FVC <70% predicted, total lung capacity (TLC) < 80% predicted or diffusing capacity of carbon monoxide (DLCO) < 80% predicted.

The recorded laboratory parameters encompassed blood hemoglobin, ferritin, C-reactive protein (CRP), interleukin-6 (IL6), N-terminal pro natriuretic peptide (NT-proBNP), D-dimer and anti-S1/S2 protein SARS-CoV2 immunoglubulin gamma (anti-S1/S2 IgG). The full list of variables with stratification scheme and procedure details are provided in Supplementary Table S1and Supplementary Methods.

### Statistical analysis

Statistical analyses were performed with R version 4.0.3 as presented in Supplementary Figure S1. Kinetics of symptom and radiological lung finding resolution were assessed with mixed-effect logistic regression^15^. Risk factor modeling was performed with fixed-effect logistic regression. Clustering of binary clinical features and of study participants was analyzed with the k-means algorithm^16^. Prediction of lung lesions by distance weighted kNN^17^ and naive Bayes^18^ algorithms was tested in 200 random training/test subset splits of the cohort data (training n = 80, test n = 28). P values were corrected for multiple comparisons by Benjamini-Hochberg method^19^, effects were termed significant for p < 0.05. Details of statistical analysis are provided in Supplementary Methods.

## Results

### Patient characteristics

The CovILD study cohort subset included in the current report (n = 108) predominately consisted of males (54.6%), and participants were aged between 19 to 87 years (Table 1). Most participants displayed preexisting comorbidity (79.6%), predominantly cardiovascular and metabolic diseases. The cohort included patients with mild (outpatient care, n = 26, 24.1%), moderate (hospitalization without oxygen supple, n = 31, 28.7%), severe (hospitalization with oxygen supply, n = 33, 30.6%), and critical (ICU treatment, n = 18, 16.7%) acute COVID-19.

**Table 1.**
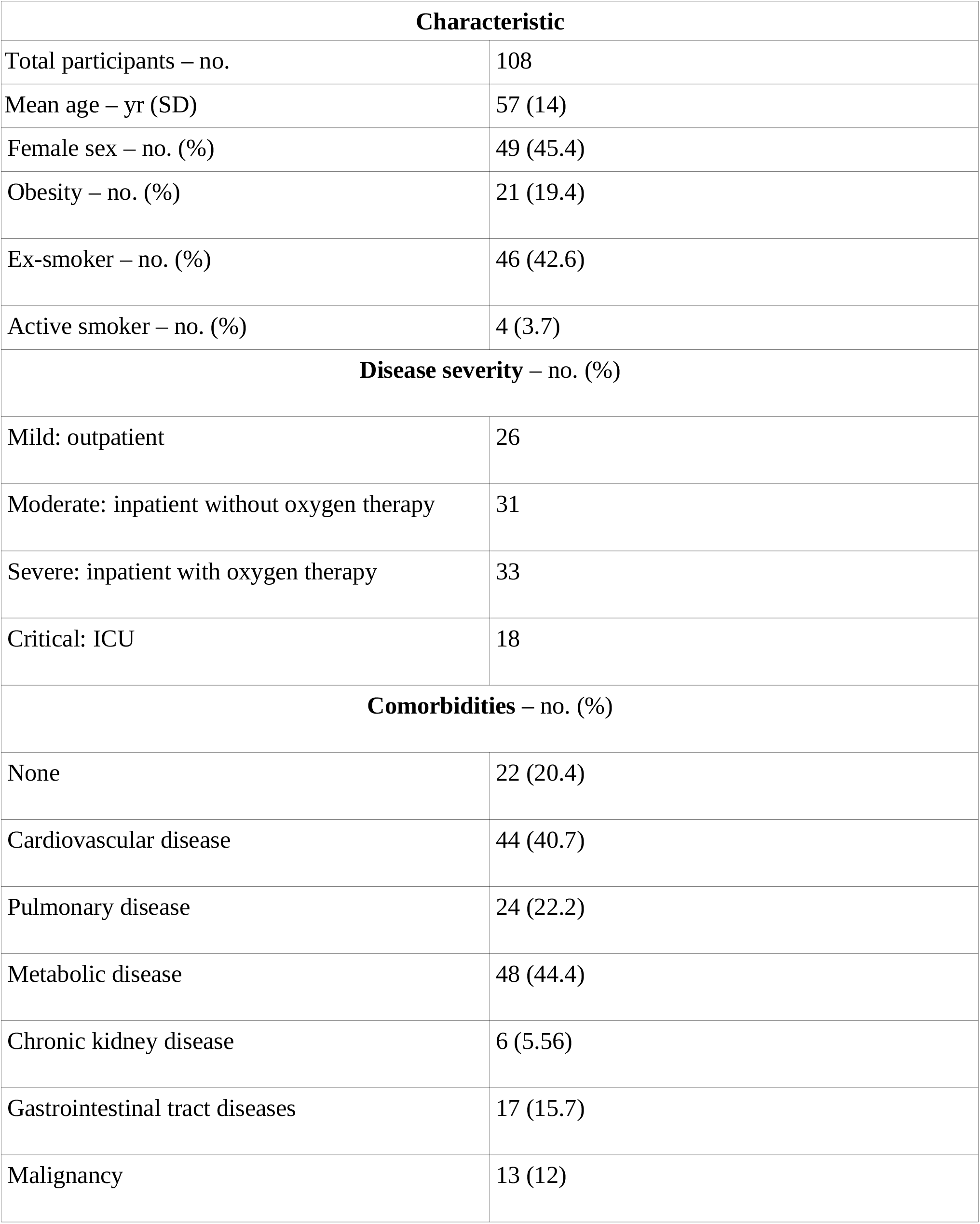
Characteristics of the study population

### Clinical recovery after COVID-19

During 180 days of COVID-19 convalescence, most patients, irrespective of the severity of the acute infection, demonstrated a significant contraction of the surveyed disease symptoms (Figure 1A). Nevertheless, 180 days after the disease onset, 51.9% of the study subjects still reported COVID-19-related complaints, with self-reported impaired physical performance (27.8%) and exertional dyspnea (24.1%) being the most frequent (Figure 1B). Prevalence of all investigated symptoms except for sleep disorders declined significantly, even though the pace of their resolution was remarkably slower in the late (100-and 180-day follow-ups) than in the early-recovery phase (till 60-day follow-up).

**Figure 1.**
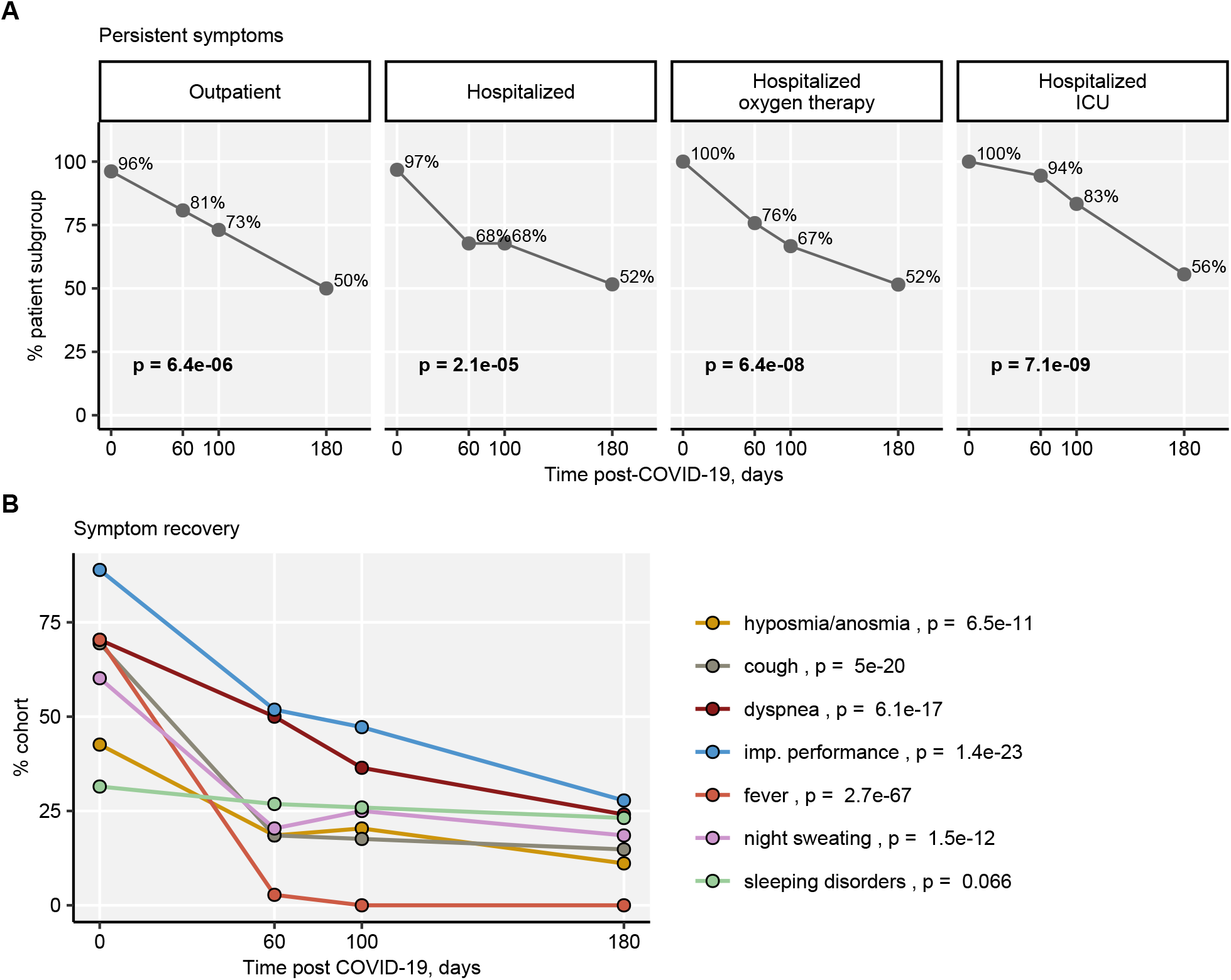
Resolution of COVID-19 symptoms. Percentages of any symptoms present in the study cohort stratified by the severity of acute disease (A) and particular symptom frequencies in the entire cohort (B) were calculated. Statistical significance was assessed by mixed-effect logistic regression and p values obtained by LRT test. In (B) separate models were fitted to each severity group. P values were corrected for multiple comparisons by Benjamini-Hochberg method. Outpatient: n = 26, hospitalized without oxygen: n = 31, hospitalized with oxygen: n = 33, ICU: n = 18, entire cohort: n = 108.

Impairment of lung function could be discerned in 33% of the entire cohort. Remarkably, except for the critical acute COVID-19 subjects (60 days: 72%, 180 days post-COVID-19: 53%), no significant reduction of the functional lung impairment prevalence was observed (Figure 2).

**Figure 2.**
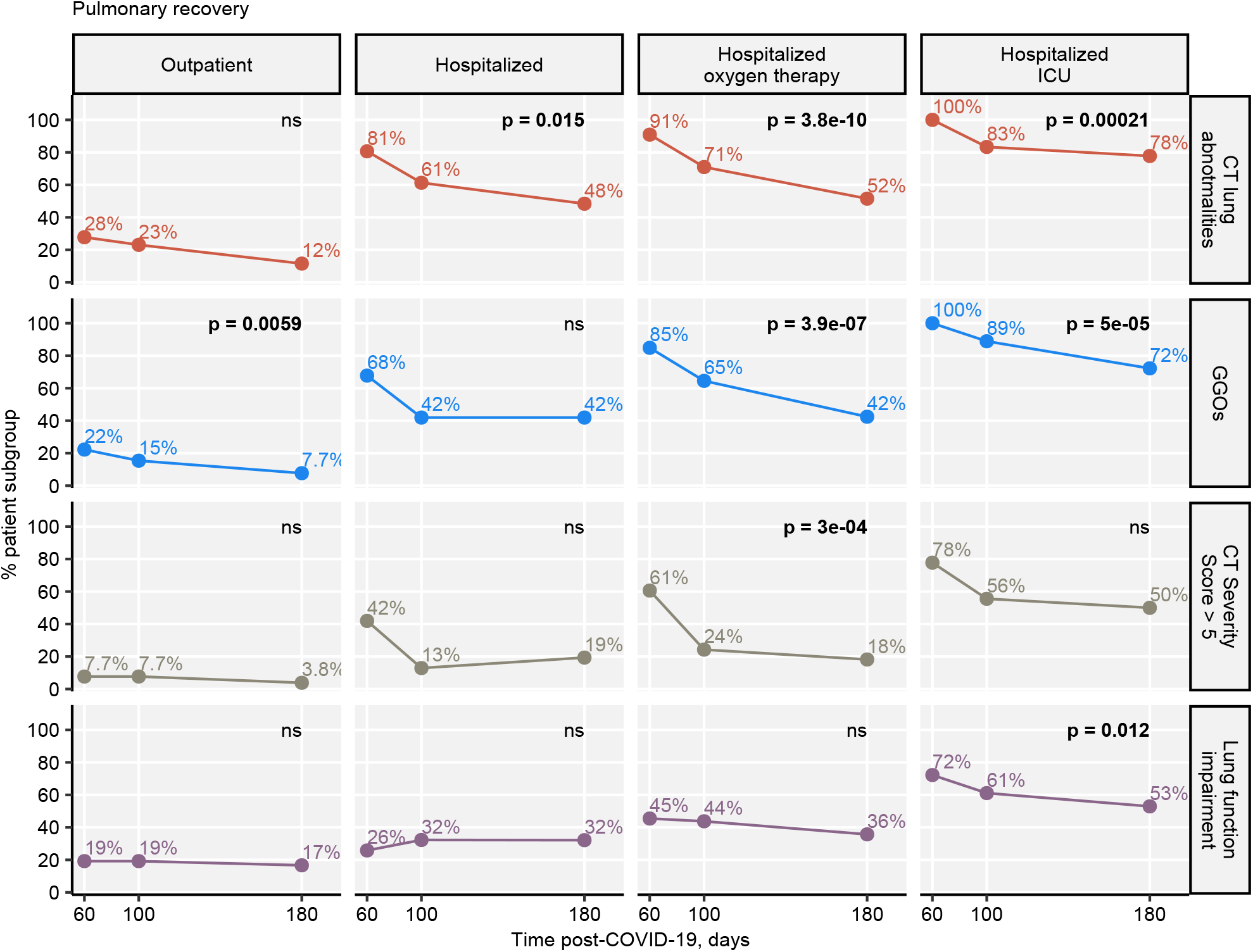
Resolution of lung lesions and functional impairment. Percentages of subjects with any lung lesions detected by CT, GGOs, CT lung abnormalities scored > five severity points and functional lung impairment in the study cohort stratified by the severity of acute disease were calculated. Statistical significance was assessed by mixed-effect logistic regression and p values obtained by LRT test. Separate models were fitted to each severity group. P values were corrected for multiple comparisons by Benjamini-Hochberg method. Outpatient: n = 26, hospitalized without oxygen: n = 31, hospitalized with oxygen: n = 33, ICU: n = 18, entire cohort: n = 108.

Abnormal structural lung findings were still found in 45.4% of patients and moderate-to-severe radiological lung alterations (CT severity score > five points) were present 20% of participants. Interestingly the radiological lung findings demonstrated only weak co-occurrence (less than 50%) with the impaired lung function at all post-COVID-19 visits (Supplementary Figure S2). As expected, the prevalence and recovery of CT lung findings were related to the severity of acute infection. The highest prevalence of any abnormalities, GGO and lesions scored above five CT severity points at the 180-day follow-up was observed in the individuals with severe and critical acute disease (Figure 2). Notably, the hospitalized group with oxygen therapy demonstrated the fastest recovery kinetics (91% and 52% subjects with any abnormalities at the 60 and 180-day visit, respectively). Furthermore, the remaining severity strata showed only a minor drop in the lung finding prevalence between the day 100 and day 180 visits, in particular regarding the moderate-to-severe pulmonary lesions (Figure 2).

### Risk factors of persistent lung lesions

To search for risk factors associated delayed pulmonary recovery, we screened a set of 50 binary demographic, clinical and biochemical parameters recorded during the acute SARS-CoV2 infection and at the 60-day visit (Supplementary Table S2) for correlation with the three readouts of persistent lung abnormalities at the six-month follow-up (Supplementary Table S2).

Among the candidate features, 22 were significantly associated with both the risk of any radiological lung abnormality and GGOs, and only eight were linked to moderate-to-severe CT detectable lesions (Supplementary Figure S2B). A total of six variables (immunosuppressive therapy, ICU stay, and over three pre-existing conditions during acute COVID-19 and CRP > 0.5 mg/L, IL6 > 7 ng/L, lung function impairment at the 60-day follow-up) were identified as the risk-modifying factors common for all three readouts (Figure 3, Supplementary Figure S2B). Among them, markers of non-resolving inflammation: elevated IL6 and elevated CRP at the early follow-up were the strongest unfavorable risk factors (Figure 3).

**Figure 3.**
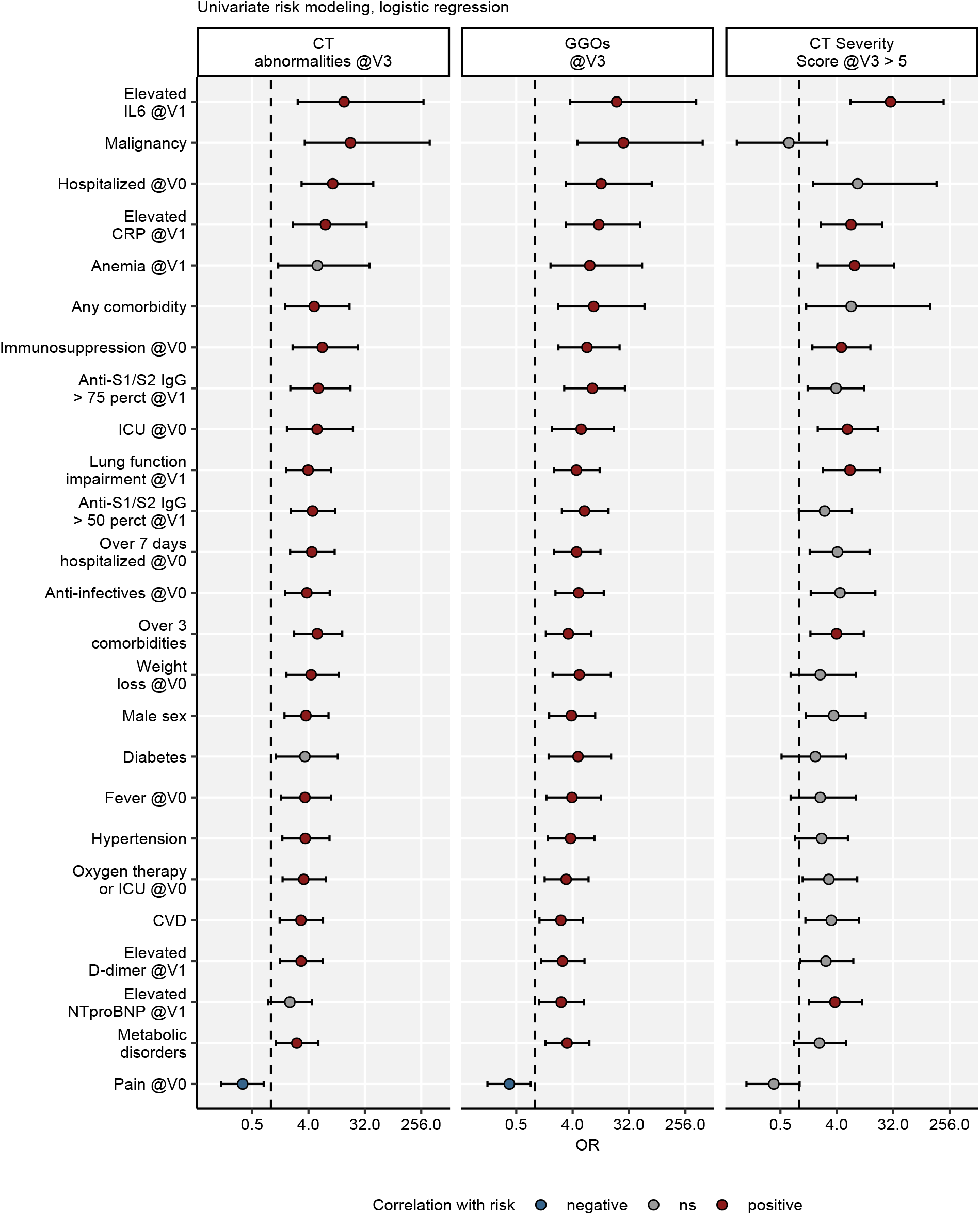
Identification of risk factors of persistent lung abnormalities at the 180-day post-COVID-19 follow-up. Correlation of candidate risk factors recorded at the disease onset of the 60-day follow-up with the presence of any radiological lung abnormality, GGOs or CT lesions graded with > five severity points at the 180-day follow-up was investigated by univariate logistic regression. Statistical significance of OR estimates was assessed by Wald Z test, p values were corrected for multiple comparisons by Benjamini-Hochberg method. Points with whiskers represent OR with 95% CI, point color codes for significance and the correlation sign. Dashed lines represent OR = 1. N = 108. V0: acute COVID-19, V1: 60-day follow-up, V3: 180-day follow-up.

### Clusters of clinical features linked to persistent lung lesions

To discern constellations of non-CT parameters linked to protracted lung recovery, we subjected the initial variable pool and the readouts of persistent lung abnormality to unsupervised clustering analysis. By this means, four clusters of clinical features were identified (Figure 4A, Supplementary Figure S3A and Supplementary Table S3).

**Figure 4.**
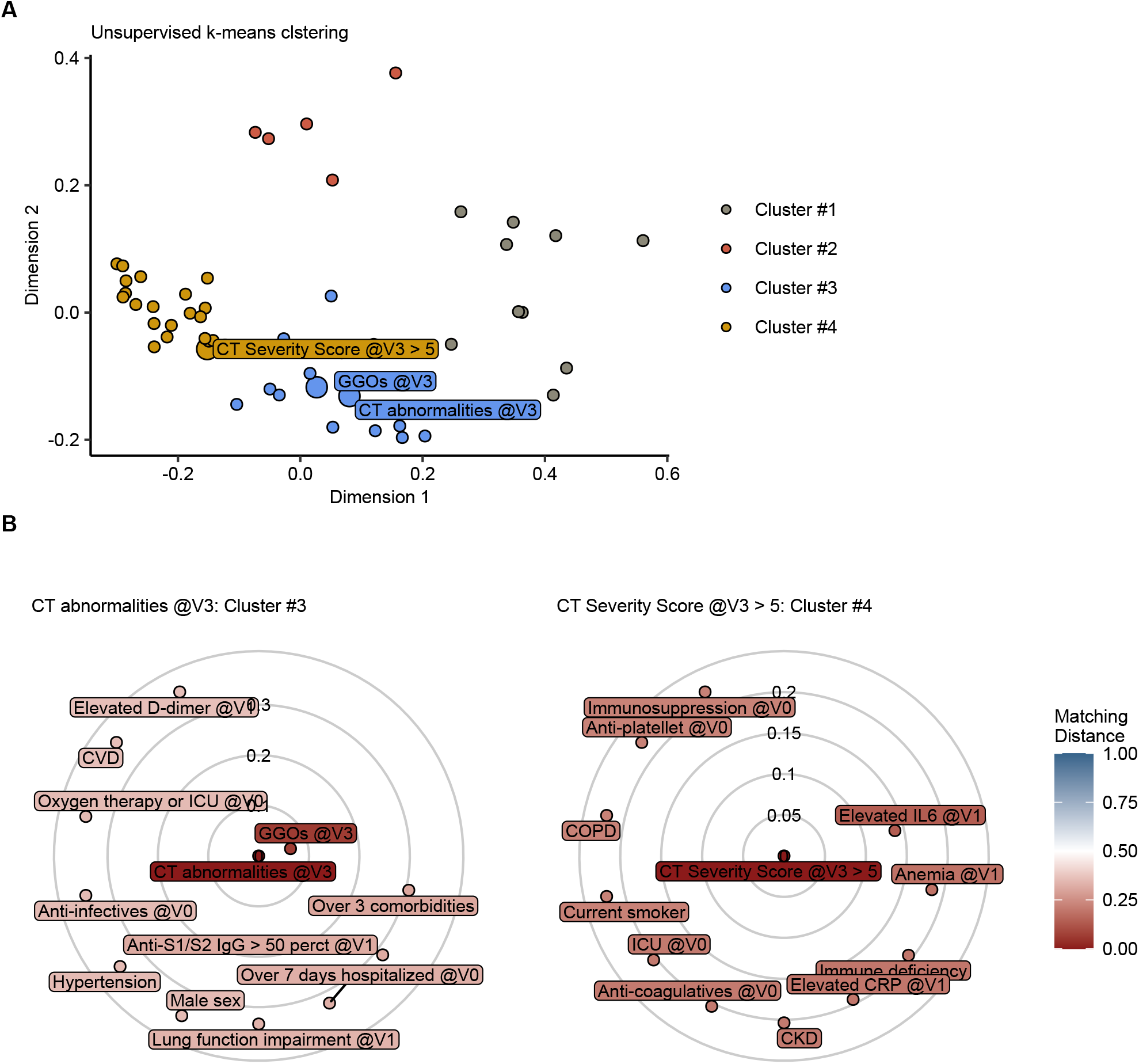
Clustering of the clinical features recorded at the disease onset, the 60-day follow-up and the lung lesion readout at the 180-day post-COVID-19 visit. Association of binary clinical features: 50 laboratory, demographic and clinical parameters at disease onset and the 60-day follow-up visit and three lung abnormality readouts (any lung abnormalities, GGOs and lesions graded > five severity points) at the 180-day post-COVID-19 visit was investigated by k-means clustering with the simple matching distance measure. V0: acute COVID-19, V1: 60-day follow-up, V3: 180-day follow-up. (A) Cluster assignment plot. For visualization, the distance matrices were subjected to two-dimensional MDS (multi-dimensional scaling). Each point represents a single clinical feature, color codes for the cluster assignment. Lung abnormality readouts were highlighted with a larger point size and labeled with their names. (B) Radial plots displaying 10 nearest cluster neighbors for any lung lesions and GGOs (cluster 3) and of CT lung lesions graded > five severity points (cluster 4). Circle radius codes for the simple matching distance from the respective lung abnormality variable.

Surprisingly, whereas any CT lung abnormality and GGOs were assigned to one common cluster (Cluster #3), CT pathology scored above five severity points was associated with a separate set of co-occurring features (Cluster #4) (Figure 4A). The ten closest cluster neighbors of any non-resolving radiological lung findings and GGOs included anti-S1/S2 IgG above the cohort median as a readout of the anti-viral immune response strength, elevated D-dimer as a marker of coagulation dysfunction and microvascular injury determined at the early follow-up together with prolonged hospitalization, oxygen and anti-infective therapy during acute COVID-19, male sex, multi-morbidity, cardiovascular disease and metabolic disorders (Figure 4B). In turn, more severe pulmonary pathology was closely linked to elevated markers of inflammation (IL6, CRP) and anemia at the early follow-up, along with hallmarks of critical severity of acute COVID-19 such as anti-coagulative/anti-platellet therapy and ICU stay. Furthermore, long-term immunosuppressive treatment, immunodeficiency, chronic kidney disease and history of smoking or COPD were also tightly linked to non-resolving moderate-to-severe CT lesions (Figure 4B).

### Automated identification of subjects at risk of perturbed pulmonary recovery

Next, we sought to define a subset of subjects at risk of the delayed pulmonary recovery with a similar unsupervised clustering procedure applied to the study participants. For clustering, we used the set of non-CT 50 clinical features used in the risk analysis and, subsequently, investigated the prevalence of lung abnormalities at the 180-day follow-up in the participant subsets.

By this approach, three sub-populations could be discerned, termed further ‘low-’, ‘intermediate-’ and ‘high-risk’ subsets (Figure 5, Supplementary Figure S3B). In sum, 23 clustering factors were significantly more frequent in the intermediate-or high-risk group than in the low-risk subsets, including primarily readouts of the anti-SARS-CoV2 immunity (anti S1/S2 IgG), disease severity (hospitalization, oxygen therapy and ICU stay, antibiotic therapy, weight loss), multi-morbidity (more than three comorbidities, CVD, hypertension, metabolic disorders), impaired lung function, age and male sex together with polysmptomatic acute COVID-19 (more than six symptoms, cough, fever). Interestingly, participants older than 65, suffering from hypercholesterolemia, males and those with functional lung impairment, elevated NT-proBNP and D-dimer levels at the 60-day follow-up were over-represented in the high-risk compared with the intermediate-risk group. In turn, polysymptomatic acute COVID-19 was more specific for the intermediate risk subset (Figure 6A).

**Figure 5.**
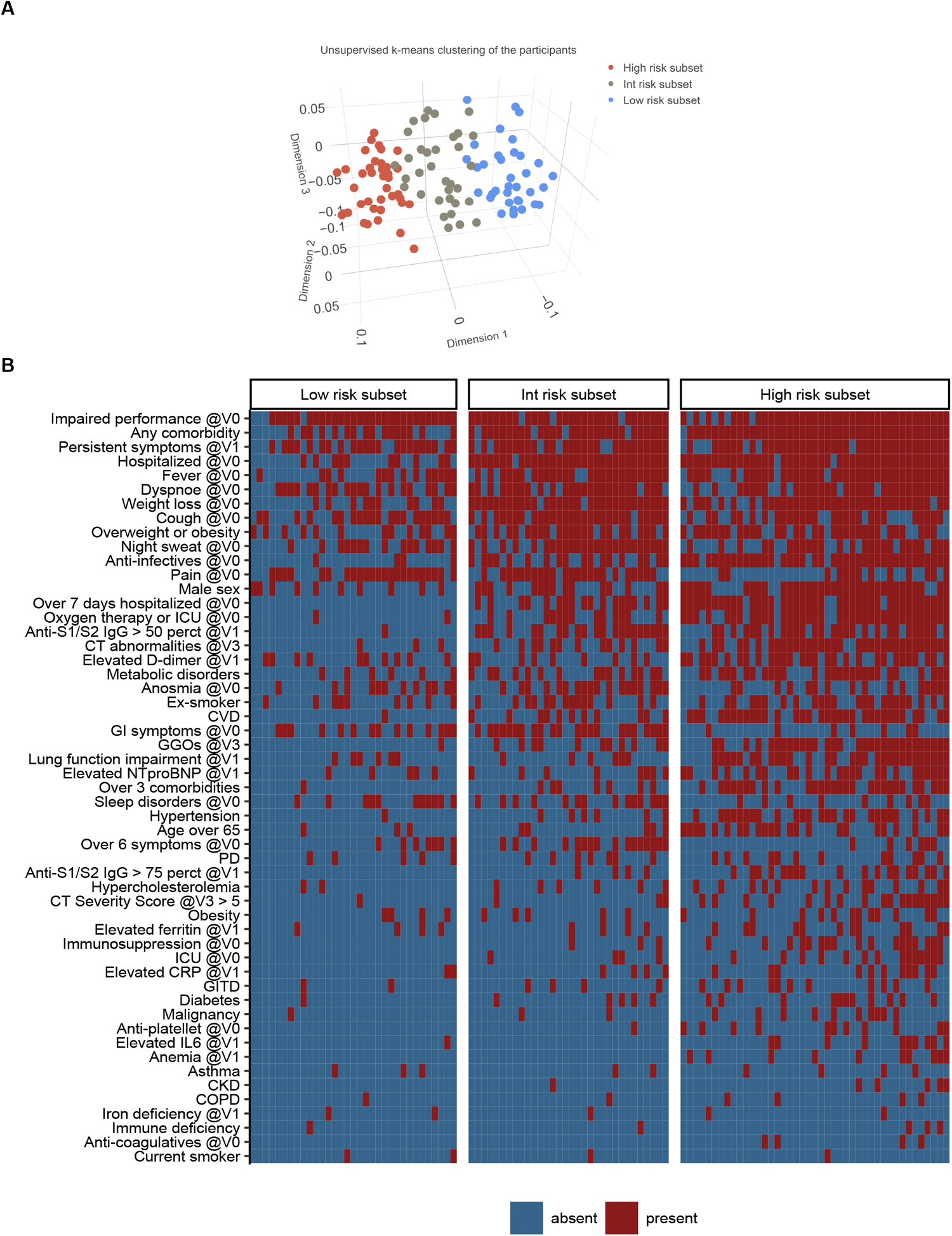
Unsupervised k-means clustering of the study participants. Study participants (n = 108) were subjected to k-means clustering in respect to 50 non-CT binary clinical features recorded at the onset and the 60-day follow-up and Jaccard distance measure. Three separate participant subsets were identified termed ‘Low-’, ‘Intermediate-’ and ‘High-Risk Subset’. V0: acute COVID-19, V1: 60-day follow-up, V3: 180-day follow-up. (A) Cluster assignment plot. For visualization, the distance matrices were subjected to three-dimensional MDS (multi-dimensional scaling). Each point represents a single study participant, color codes for the cluster assignment. (B) Presence or absence of the clustering features in the participants assigned to the low, intermediate and high risk subsets.

**Figure 6.**
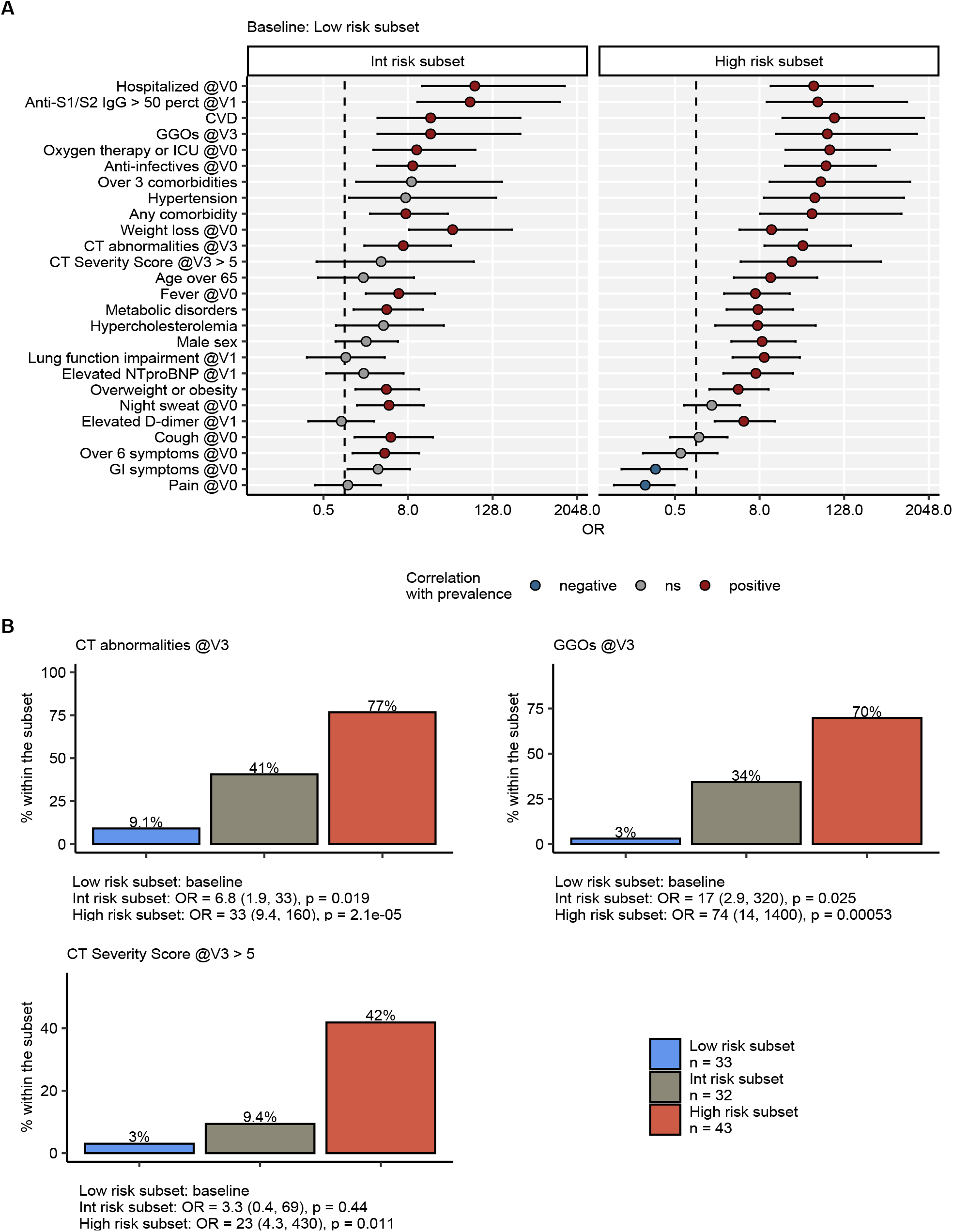
Prevalence of non-CT clinical features and readouts of long-term radiological lung abnormality in the low-, intermediate-and high-risk study subject subsets. Study subjects were assigned to the low-, intermediate-and high-risk subsets by k-means clustering as presented in Figure 5. Differences in prevalence of clinical features between the intermediate-or high-risk subset and the low-risk subset were modeled with logistic regression. Statistical significance of OR estimates was assessed by Wald Z test, p values were corrected for multiple comparisons by Benjamini-Hochberg method. V0: acute COVID-19, V1: 60-day follow-up, V3: 180-day follow-up. A) Points with whiskers represent OR with 95% CI, point color codes for significance and the correlation sign. Dashed lines represent OR = 1. (B) Prevalence of any lung abnormalities, GGOs and lesions graded > five severity points in CT at the 180-day follow-up in the low, intermediate and high-risk subsets.

Most importantly, any persistent CT lung abnormalities and GGO were significantly more prevalent in the intermediate-and high-risk that in the low-risk subset (Figure 6B). In turn, CT lung lesions scored more than five severity points were significantly enriched only in the high-risk subset and displayed comparable low prevalence in the remaining sub-populations (Figure 6B). Because of the implementation of the disease severity variables in the participant clustering procedure, the fraction of intermediate and high risk subset cases grew with the severity of acute COVID-19 (Supplementary Figure S4A). However, adjustment of the risk modeling for the hospitalization/ventilation and ICU status had no substantial effect on the prediction of persistent lung abnormalities by the risk subset assignment (Supplementary Figure S4B).

Finally, given the significant differences in pulmonary recovery between the participant risk subsets, we asked if the long-term lung abnormality could be reliably predicted based solely on the non-CT parameters available till the 60-day follow-up. To this end, we applied two simple machine learning procedures, k-nearest neighbor (kNN)^17^ and naive Bayes algorithm^18^ to the pool of non-CT 50 binary clinical features and multiple random splits of the study cohort into the training and test data sets. This approach differentiated between the complete pulmonary recovery and presence of any CT lung lesions at the 180-day follow-up with an accuracy exceeding 70% (kNN: 71% for kNN, naive Bayes: 75%) and sensitivity ranging from 63% (naive Bayes) to 75% (kNN). Of note, similar prediction quality was achieved at detection of persistent GGOs by both tested procedures. In contrast, only the naive Bayes algorithm succeeded at predicting the less frequent moderate-to-severe lung lesions (accuracy: 64%, sensitivity: 71%) whereas the kNN procedure failed to identify the majority of them (sensitivity: 33%) (Figure 7, Supplementary Table S4). Importantly, the investigated procedures efficiently identified the non-resolving pulmonary findings both in the mild-to-moderate (outpatients and inpatients without oxygen) and severe-to-critical (ventilated inpatients and ICU) participant subsets, even though the specificity varied between the procedures and CT abnormality readouts (Supplementary Figure S5 and S6).

**Figure 7.**
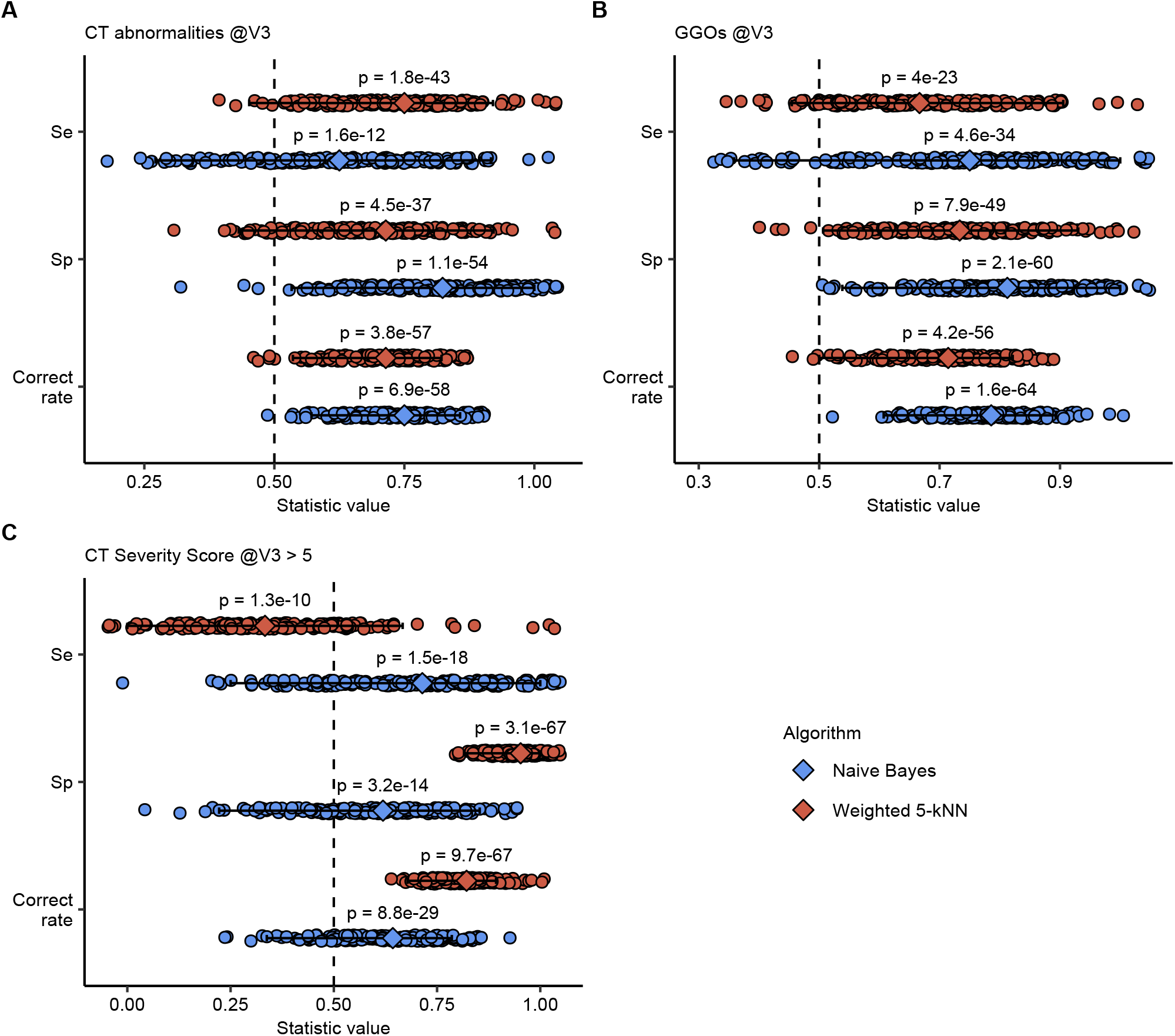
Prediction of persistent lung lesions based on acute COVID-19 and early follow-up non-CT variables by the kNN and naive Bayes machine learning algorithms. Ability to predict any CT lung lesions, GGOs and lesions graded > five severity score at the 180-day follw-up (V3) based on the set of 50 non-CT binary clinical parameters recorded during acute COVID-19 and the 60-day follow-up was tested with the distance-weighted k-nearest neighbors (kNN) (k = 5, Jaccard distance between the subjects, random tie resolution) and naive Bayes algorithms. Correct prediction rate, sensitivity and specificity of the algorithm were assessed with 200 random training/test splits of the initial data set (training: n = 80, test = 28). The significance of the correct prediction rates, sensitivity and specificity versus random predictions was determined by the Mann-Whitney U test. Each point represents a single training/test split analyzed, diamonds with whiskers represent expected values (median), 2.5% and 97.5% percentile of the statistic.

## Discussion

Over a year of the pandemic, the long-term trajectories of COVID-19 recovery and their individual variability are still poorly characterized^10^. Recent survey studies suggest that post-COVID-19 syndrome defined as persistence of symptoms for at least twelve weeks may affect as many as 10% of COVID-19 patients^20–22^. These potentially massive numbers of post-COVID-19 sequelae pose an enormous socioeconomic and medical challenge. Hence, robust, evidence-based tools for their prediction, monitoring and treatment are urgently needed^12,23^. Herein, we longitudinally investigated a cross-sectional cohort of COVID-19 convalescents differing in demographic and clinical features and in severity of the acute infection and prospectively assessed the course of symptom and pulmonary recovery.

A reported previously^5,10^, we could discern a measurable to complete pulmonary recovery assessed by lung function or CT at the six-month follow-up even if the acute disease was severe or critical. However, persistent COVID-19 symptoms and structural lung abnormalities were detected in more than 40% and reduced lung function in approximately one-third of the participants. Furthermore, we could observe a deceleration of the pulmonary and symptom recovery between the three-and six-month assessments, which may point toward chronicity of the post-COVID-19 pulmonary damage. Notably, such prolonged structural and functional lung recovery in a time window of two to five years after acute disease was reported for SARS^9,11^ and non-COVID-19 acute respiratory distress syndrome (ARDS)^24,25^.

Even though severe or critical acute COVID-19 is regarded as the major indicator of an unfavorable long-term outcome^10,23,26^, it is not helpful at predicting complications of pulmonary recovery from mild or moderate infection. Our systematic search of risk factors in a pool of clinical variables recorded during acute COVID-19 and at the two-month follow-up revealed, apart from the acute disease severity, multi-morbidity and male sex reported by us previously^5^, markers of elevated inflammation, tissue/vascular damage and anti-SARS-CoV2 immunity at the early post-COVID-19 assessment. In particular, elevated blood CRP and IL6 were tightly correlated with all investigated CT lung abnormalities, suggesting the contribution of non-resolving pathological inflammation to the persistence of pulmonary lesions. Notably, only a minor overlap with the radiological lung findings and reduced lung function at the consecutive follow-up assessments was observed. Hence the sole application of a lung function measurement at screening for subjects at risk of delayed lung recovery may bear insufficient sensitivity. In turn, adjunct monitoring of standard inflammatory parameters such as IL6 or CRP analogous to systemic sclerosis^27^ may greatly augment identification of the individuals at risk of chronicity of pulmonary damage. At the same time, our observation points to the necessity of further research on the inflammatory background of the post-COVID lung damage and infers a potential benefit of anti-inflammatory therapy. Surprisingly, except of fever during acute COVID-19, none of the surveyed symptoms, symptom burden or persistence could be linked to the long-term lung injury risk. This is in an apparent contrast to the generalized, predominantly mild-course COVID-19 convalescent population where patterns of symptoms patterns and burden during acute infection were described as long-COVID predictors^20^.

Employing an in-depth clustering, we could in addition observe distinct patterns of clinical features co-occurring with any CT lung abnormality, GGOs and moderate-to-severe lung lesions at the six-month follow-up. Interestingly, high anti-S1/S2 IgG and D-dimer levels at the two-month follow-up but not the inflammatory markers were closely associated with the first two abnormality readouts. This let us speculate that the magnitude of the adaptive anti-viral immunity and organ damage without systemic inflammatory background are drivers of mild and moderate long-term lung abnormalities frequently observed in our cohort. In turn, CT lesions graded over five severity points were primarily associated with elevated IL6, CRP and inflammatory anemia^28^ during early convalescence, smoking and COPD. Thus, genesis and persistence of less frequent moderate-to-severe pulmonary lesions may additionally require an interplay between strong, prolonged inflammation and pre-existing lung injury.

With a similar classification technique based on non-CT clinical features of acute COVID and early recovery we could characterize three sub-populations of convalescents significantly differing in the overall prevalence and severity of CT lung findings at the six-month follow-up. Importantly, although multiple readouts of the COVID-19 severity were implicitly included in the clustering algorithm, the intermediate-or high-risk subset assignment remained strongly predictive of long-term lung abnormality even upon adjustment for the hospitalization/ventilation and ICU status. This underlines further the vital importance of the parameters not directly connected to the severity of acute infection, e. g. ongoing inflammation or pre-existing lung injury for the comprehensive risk assessment.

Finally, in addition to the unsupervised clustering, we demonstrate the utility of two technically unrelated machine learning procedures, kNN^17^ and naive Bayes^18^, at assessing the individual risk of a perturbed recovery based solely on non-CT readouts available till the two-month follow-up. Despite lacking optimization of the variable pool and small study cohort sub-populations as model training sets, high prediction correctness was achieved for any CT lung findings and GGOs at the 180-day visit by both procedures. In turn, only the naive Bayes algorithm inherently more sensitive towards rare events succeeded at identification of the less frequent moderate-to severe lesions. In clinical practice, such automated procedures provided with a larger multi-center training data set may pose inexpensive, fast and reliable tools for screening e. g. medical records for the COVID-19 convalescents at risk of poor pulmonary recovery requiring a denser follow-up and lung imaging. Recently, a similar approach was proposed for monitoring COVID-19 patients for the need of respiratory support^29^.

Our study bears limitations primarily concerning the low sample size and the cross-sectional character of the trial. Furthermore, data incompleteness and selection bias linked to disease severity (e. g. mild cases were not subjected to CT scans during acute COVID-19) resulted in a considerable dropout rate and potentially confounded the clustering and risk prediction analyses. Additionally, the candidate risk factors and the risk-assessment algorithms of perturbed pulmonary recovery presented here call for verification in a larger, independent multi-center collective of COVID-19 convalescents.

In summary, we herein present a comprehensive description of the resolution of symptoms and structural pulmonary abnormalities in the first 6 months of COVID-19 convalescence. We report a high frequency of lung abnormalities and symptoms present in almost half of the studied population and a flattened recovery kinetics after three-months post-COVID-19. Systematic risk modeling and clustering analysis reveled a set of clinical variables linked to protracted recovery apart from the severity of acute infection such as inflammatory markers, anti-S1/S2 IgG, multi-morbidity, and male sex. Of practical importance, we demonstrate that automated classification algorithms may help to identify individuals at risk of persistent lung lesions and relocate resources to prevent long-term disability.

## Data Availability

As the longitudinal data presented in the manuscript refer to an ongoing study, the complete data set will be made available as soon as the study is completed.

## Author Contribution

T.S., I.T., and J.LR. designed the study. S.S., A.B., M.A., T.S., A.P., A.L., C.S., K.K., S.K., M.N., B.P., A.E, G.H., E.W., J.LR., I.T. performed the clinical investigations and collected the data. T.S. and P.T. performed data analysis. T.S., P.T., A.L., C.S., G.W., G.We., I.T., and J.LR. interpreted the data. T.S., P.T., J.LR., and I.T. wrote the manuscript. All authors critically reviewed the final version of the manuscript.

## Acknowledgments

We acknowledge the commitment of the staff and providers of our institutions through the COVID-19 crisis and the suffering and loss of our patients as well as their families.

## Abbreviations

ARDS: acute respiratory distress syndrome
SARS: severe acute respiratory syndrome
CI: confidence interval
COVID-19: coronavirus disease
2019 CRP: C-reactive protein
CT: computed tomography
CVD: cardiovascular disease
GGO: ground-glass opacity
IL6: interleukin-6
kNN: k nearest neighbors
LRT: likelihood ratio test
NT-proBNP: N-terminal pro brain natriuretic peptide
OR: odds ratio
SARS-CoV2: severe acute respiratory syndrome coronavirus 2

