## Supplementary Material for "Investigating phenotypes of pulmonary COVID-19 recovery – a longitudinal observational prospective multicenter trial"

### Supplementary Methods

#### Procedures

We retrospectively assessed patient characteristics during acute COVID-19 and performed follow-up investigations at 60 days (63 ± 23 days (mean ± SD); visit 1), 100 days (103 ± 21); visit 2) and 180 days (190 ± 15; visit 3) after the diagnosis of COVID-19. Each visit included clinical examination, assessment of typical COVID-19 symptoms and performance status with a standardized questionnaire, lung function testing, capillary blood gas analysis, trans-thoracic echocardiography, standard laboratory testing and a low-dose computed tomography (CT) scan of the chest. The full list of queried variables with cutoffs and stratification scheme is presented in Supplementary Table S1.

Serological markers were determined in EDTA blood with standardized procedures at the Rheumatology and Infectious Diseases Laboratory (RILA) and the Central Institute of Clinical and Chemical Laboratory Diagnostics  (ZIMCL) of the University Hospital of Innsbruck. C-reactive protein (CRP),  interleukin-6 (IL6), N-terminal pro natriuretic peptide (NT-proBNP), and serum ferritin were measured using a Roche Cobas 8000 analyzer. D-dimer was determined with a Siemens BCS-XP instrument using the Siemens D-Dimer Innovance® reagent (D-dimer). Anti-S1/S2 protein SARS-CoV2 immunoglubulin gamma were quantified with LIAISON® SARS-CoV-2 S1/S2 IgG chemoluminescence assay (CLIA, DiaSorin, Italy) and expressed as arbitrary units. For the analysis, antibody levels in the cohort were stratified by concentration quartiles.

Low dose (100 kVp tube potential) CT scans of the chest were acquired in craniocaudal direction without iodine contrast agent without ECG gating on a 128 slice multidetector CT with a 128×0.6 mm collimation and spiral pitch factor of 1.1 (SOMATOM Definition Flash, Siemens Healthineers, Erlangen, Germany). In case of the clinical suspicion for pulmonary embolism (PE), CT scans were performed with a contrast agent. A slice thickness of 1 mm was used for axial reconstructions. CT scans were evaluated for the presence of ground-glass opacities (GGO), consolidations, bronchial dilation, and reticulations as defined by the Fleischner society. Lung findings were graded with a quantitative CT severity score (0-25 points), as previously published[1].

Lung function was deemed impaired when at least one of the following criteria was met: (1) forced vital capacity (FVC) < 80% predicted, (2) forced expiratory volume in 1 second

($FEV_1$) < 80% predicted, FEV1:FVC <70% predicted, total lung capacity (TLC) < 80% predicted or diffusing capacity of carbon monoxide (DLCO) < 80% predicted.

#### Data transformation and visualization

Demographic, clinical, biochemical, imaging, and self-reported data from the CovILD study[2] was analyzed and visualized with R programming suite (version 4.0.3). General data transformation tasks were accomplished by *tidyverse* environment[3]. Graphical visualization of the data and modeling results was done with packages *ggplot2*[4], *plotly*, *plotROC*[5] and *cowplot*[6].

An extensive follow-up assessment was performed at 60, 100, and 180 days after COVID-19 diagnosis. The recorded variables were re-coded and stratified used widely accepted cutoffs as presented in Supplementary Table S1. To account for the severity of the acute infection, study participants were classified as outpatients, hospitalized patients without oxygen or ICU need, hospitalized individuals with oxygen therapy, and ICU patients as described in our recent report[1]. In the final analyses a collective of 108 CovILD participants with a complete data record was included (Supplementary Figure S1).

#### Modeling kinetics of symptom and lung lesion recovery

To model symptom, CT lung findings and functional impairment coded as binary variables mixed-effect logistic regression was applied (random effect: individual, fixed effect: time, packages *lme4*[7] and *lmerTest*[8]). For analyses with stratification by severity groups, separate models were applied to each patient subset. Significance of symptom or radiological lung recovery was assessed by likelihood ratio test (LRT) against the respective random-term-only model.

#### Risk modeling

To identify factors associated with risk of persistent CT lung abnormality, ground glass opacities (GGO) and intermediate – high grade lung lesions (severity score > five) at the 180-day follow-up a series of uni-variate fixed-effect logistic models was generated, one for each binary modeling variable listed in Supplementary Table S1. The results were presented as odds ratio (OR) with 95% confidence intervals. The significance of the correlation (p(OR ≠ 1)) was determined by Wald Z test. P values were corrected for multiple comparisons with

Benjamini-Hochberg/FDR method[9]. An analogical modeling approach was used to determine feature prevalence differences between the low, intermediate and high risk participant subsets.

#### Cluster analyses

For clustering of 53 binary clinical features (50 clinical parameters and three CT lung abnormality readouts) simple matching distances between the features were calculated using *smc()* function from *scrime* package. For clustering of 108 study participants by 50 binary clinical features without radiological readouts, Jaccard distances between the subject were calculated using *distance()* function from *philentropy* package[10]. Both distance matrices were subjected to clustering with *kmeans()* function of base R and Hartigan-Wong algorithm[11]. The optimal number of centers (k = 4 for the features and k = 3 for the subjects) was determined by the 'elbow' method with plots of total within-cluster sum of square versus center number and package *factoextra* (function *fviz_nbclust()*). For visualization of the cluster assignment structure, the distance matrices were subjected to two- (features) or three-dimensional (subjects) MDS (multi-dimensional scaling, function *cmdscale()*, base R).

#### Prediction of lung abnormalities by the kNN and naive Bayes algorithms

The ability of the k-nearest neighbors (kNN)[12] and naive Bayes procedure[13] to predict non-resolving CT lung abnormalities (any abnormality, GGO and CT severity score > 5) at the 180-day follow-up based on 50 binary variables (without CT readouts) collected at the disease onset and the 60-day follow-up was tested with 200 random training/test subset splits of the initial data set (training: n = 80, test: n = 28). The kNN predictions were made with a home-developed distance-weighted kNN function, k = 5, Jaccard distance measure between the subjects, $distance^{-1}$ kernel function and random tie resolution. The optimal k parameter and the kernel function combination was chosen by serial comparison of prediction accuracy (correct prediction rate) with an identical set of 50 random training/test data sets. The naive Bayes predictions were made with the *naiveBayes()* function provided by *e1071* package. Prediction accuracy (error and correct prediction rate, sensitivity, specificity) was assessed by comparing the predicted outcome with the real outcome in the test sets. Confidence intervals of the prediction quality statistics were determined by BCA algorithm[14]. The significance of the calculated prediction accuracy measures was determined by comparing them with the random predictions generated for each training/test data split with Mann-Whitney U test. The tools used for prediction and accuracy testing are publicly available from GitHub (https://github.com/PiotrTymoszuk/kNN).

#### Supplementary Table S1

Variables collected during acute COVID-19 (V0) at the study visits (V1: 60-day follow-up, V2: 100-day follow-up, V3: 180-day follow-up) and analyzed in the report. The table is available online.

#### Supplementary Table S2

Identification of factors associated with the risk of any lung abnormality, GGOs or lesions graded with > 5 points in CT at the 180-day follow-up (V3) by uni-variate logistic regression. Variable: candidate risk factor, Response: CT lung abnormality readout, OR: odds ratio, 2.5% CI and 97.5% CI: bottom and top limits of the 95% confidence interval of OR, raw p: uncorrected p values obtained by Wald Z test, pFDR: raw p corrected for multiple comparisons by Benjamini-Hochberg/FDR method. The table is available online.

#### Supplementary Table S3

Cluster assignment of the 50 investigated clinical features. V1: 60-day follow-up, V3: 180-day follow-up.

| Cluster | Features |
|---|---|
| Cluster #1 | Weight loss during acute disease, Dyspnoe during acute disease, Cough during acute disease, Fever during acute disease, Night sweat during acute disease, Impaired performance during acute disease, Any comorbidity, Overweight or obesity during acute disease, Persistent symptoms @V1, Hospitalized during acute disease |
| Cluster #2 | Pain during acute disease, GI symptoms during acute disease, Anosmia during acute disease, Sleep disorders during acute disease, Over 6 symptoms during acute disease |
| Cluster #3 | CT abnormalities @V3, GGO @V3, Male sex, Ex-smoker, CVD, Hypertension, Metabolic disorders, Anti-infectives during acute disease, Elevated NTproBNP @V1, Elevated D-dimer @V1, Lung function impairment @V1, Age over 65, Over 7 days hospitalized during acute disease, Over 3 comorbidities, Anti-SARSCov2 > 50 perct @V1, Oxygen therapy or ICU during acute disease |
| Cluster #4 | CT Severity Score @V3 > 5, Obesity, Current smoker, PD, COPD, Asthma, Hypercholesterolemia, Diabetes, CKD, GITD, Malignancy, Immune deficiency, Anti-platellet during acute disease, Anti-coagulatives during acute disease, Immunosuppression during acute disease, Anemia @V1, Elevated ferritin @V1, Elevated CRP @V1, Elevated IL6 @V1, Iron deficiency @V1, Anti-SARSCov2 >75 perct @V1, ICU during acute disease |

#### Supplementary Table S4

Accuracy of prediction of radiological lung lesions at the 180 day follow-up (any CT lung abnormality, GGO and lesions graded > five CT severity points) by the distance-weighted kNN and naive Bayes algorithms. Statistic values with 95% confidence intervals are shown.

| Algorithm | | **Any CT abnormality** | **GGO** | **CT lesions > 5 severity points** |
|---|---|---|---|---|
| kNN | Sensitivity | 0.75 [0.45 to 0.92] | 0.67 [0.45 to 0.91] | 0.33 [0 to 0.67] |
| | Specificity | 0.71 [0.43 to 0.92] | 0.73 [0.51 to 0.93] | 0.95 [0.82 to 1] |
| | Correct prediction rate | 0.71 [0.54 to 0.82] | 0.71 [0.5 to 0.82] | 0.82 [0.68 to 0.89] |
| Naive Bayes | Sensitivity | 0.62 [0.27 to 0.92] | 0.75 [0.36 to 1] | 0.71 [0.25 to 1] |
| | Specificity | 0.82 [0.53 to 1] | 0.81 [0.54 to 1] | 0.62 [0.22 to 0.85] |
| | Correct prediction rate | 0.75 [0.57 to 0.86] | 0.79 [0.61 to 0.89] | 0.64 [0.34 to 0.79] |

### Supplementary Figure S1

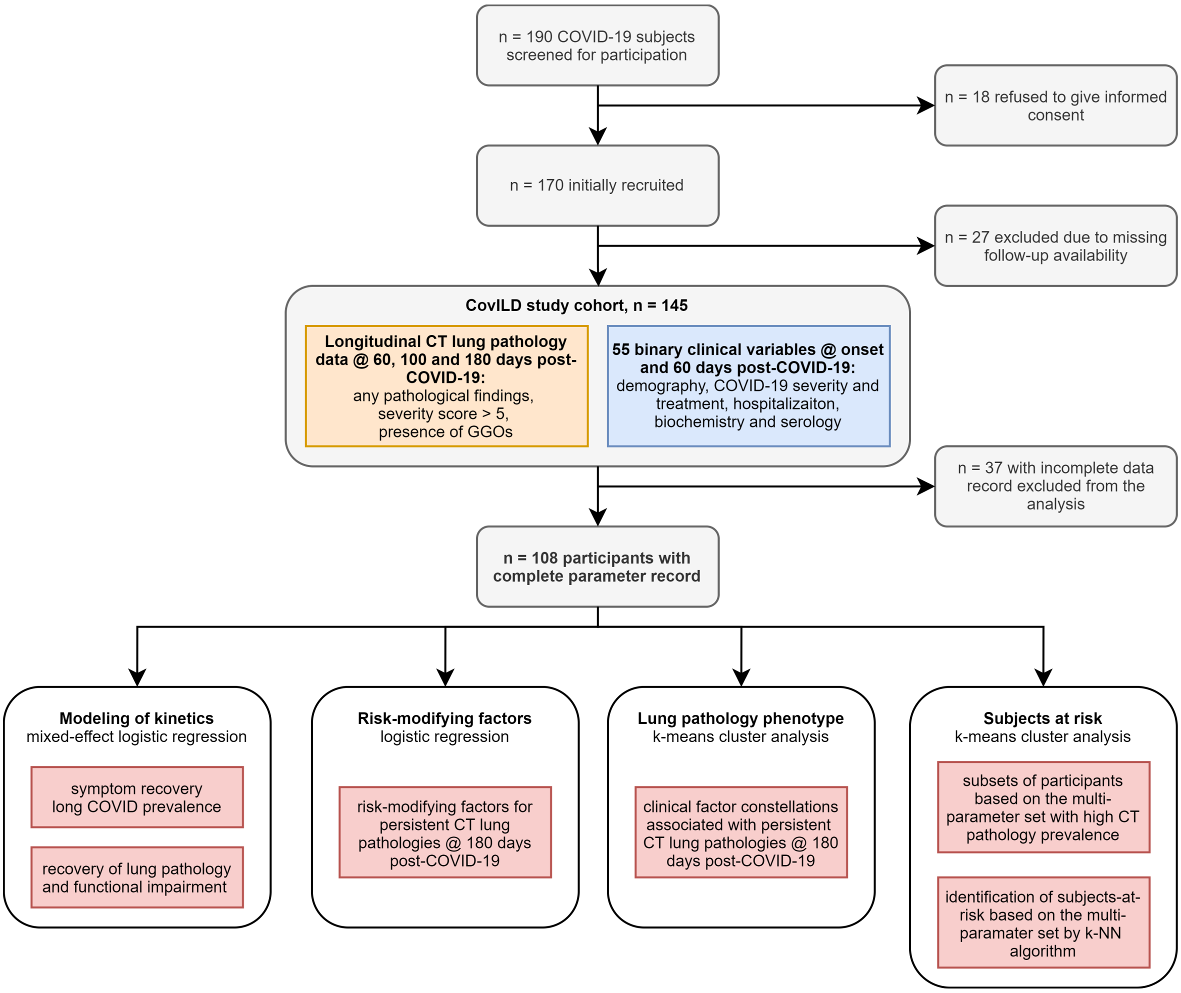


**Supplementary Figure S1. Scheme of participant screening and enrollment and data analysis.**

### Supplementary Figure S2

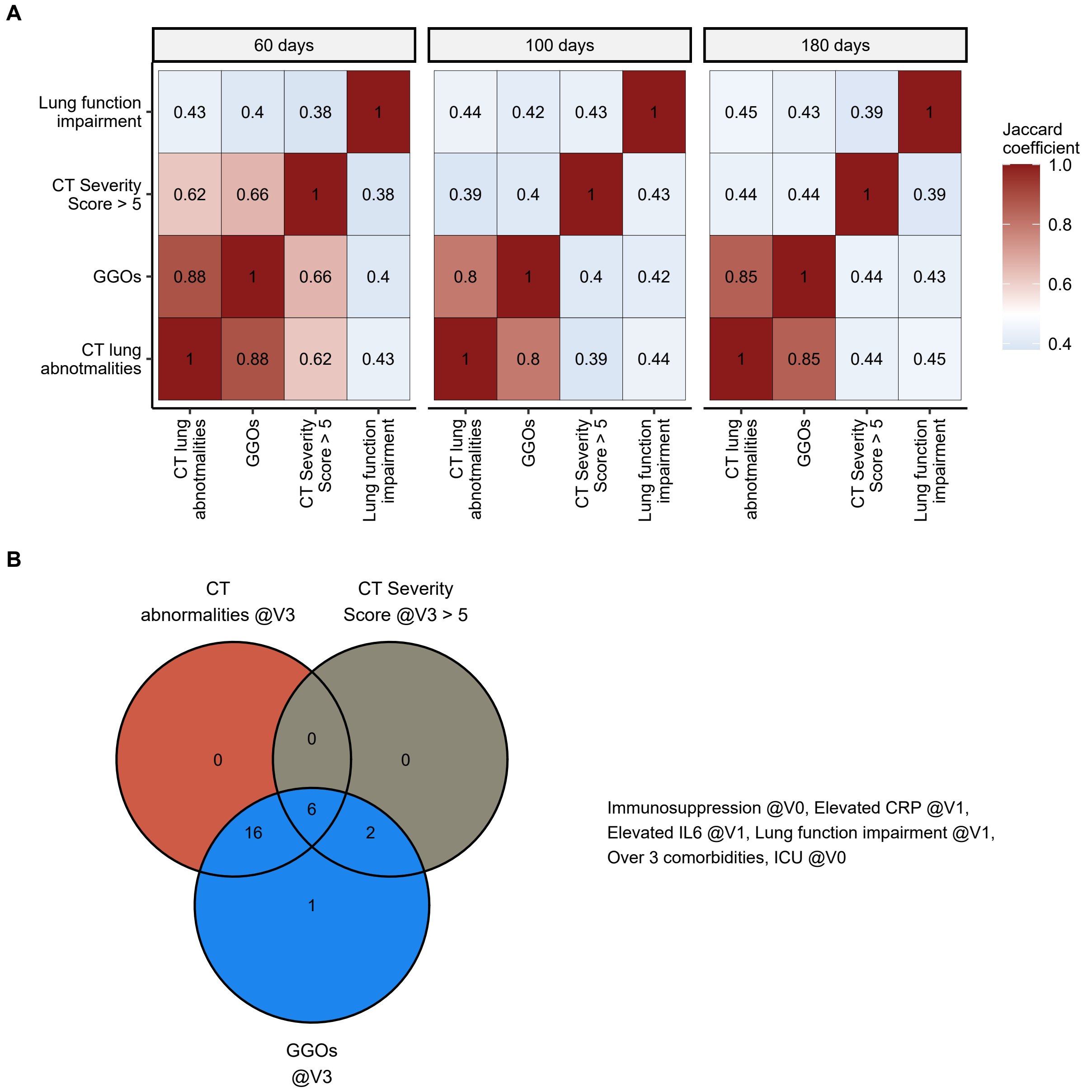


**Supplementary Figure S2. Co-occurrence of lung functional impairment and radiological lung findings in COVID-19 convalescents. Common factors associated with non-resolving lung lesions.**

(A) Co-occurrence of any lung lesions detected by CT, presence of GGO, CT severity score above >5 and impairment of pulmonary function was assessed by Jaccard coefficient (0 no co-occurrence, 1 100% overlap between the features) at the 60-, 100- and 180-day follow-up visit.

(B) Factors significantly associated with risk of any lung lesions detected by CT, presence of GGOs and lesions graded > 5 points of CT severity score at the 180-day post-COVID-19 visit were identified by uni-variate logistic modeling. Number of the significant factors shared between the CT lung abnormality responses were presented in the Venn plot. The factors common for all three responses are listed next to the plot. V0: acute COVID-19, V1: 60-day follow-up, V3: 180-day follow-up.

### Supplementary Figure S3

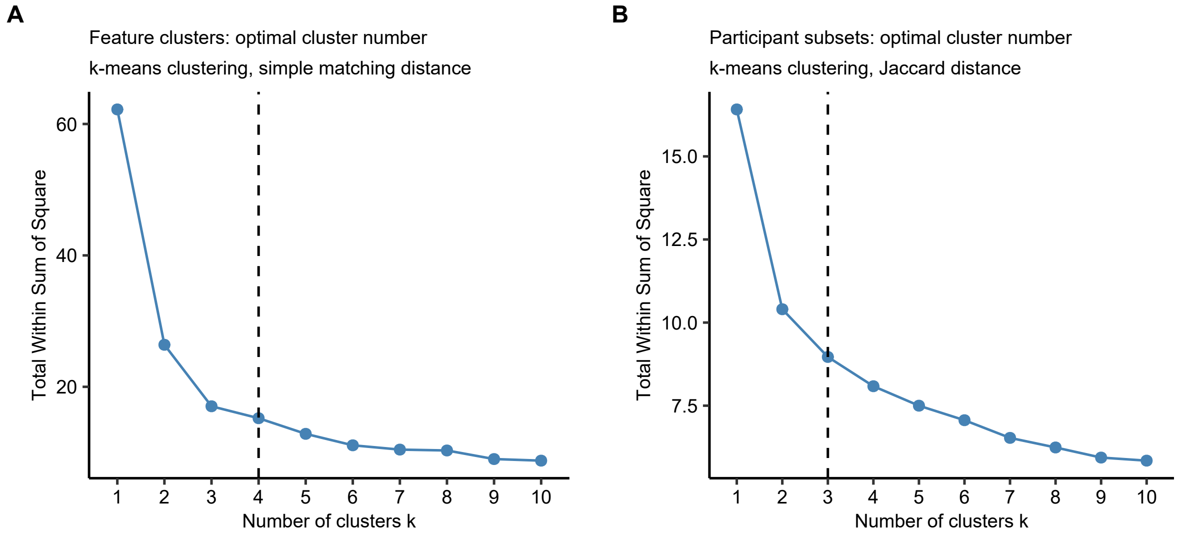


**Figure S3. Determination of the optimal cluster number in the clustering analysis of clinical features and study subjects.**

To determine the optimal cluster number, the 'elbow' method was applied based on the plot of total within-cluster sum of square values against the cluster number k for analysis of the clinical features (A) and study subjects (B).

#### Supplementary Figure S4

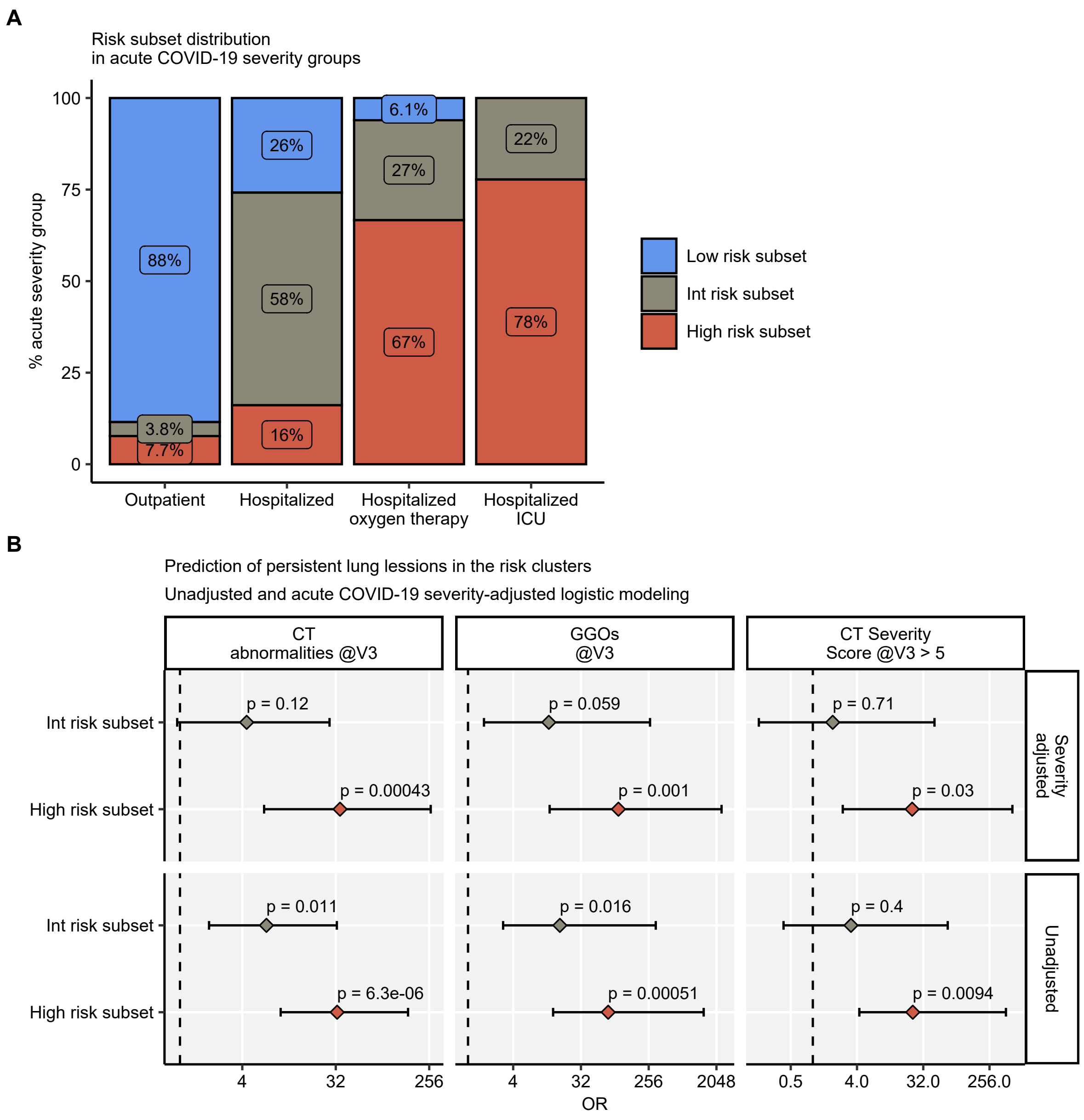


**Figure S4. Risk of persisting radiological lung damage at the 180 day follow-up in the low, intermediate and high risk subsets in study participants stratified by acute COVID-19 severity.**

(A) Assignment of acute COVID-19 outpatients (n = 26), inpatients without oxygen treatment (n = 31), ventilated inpatients (n = 33) and ICU patients (n = 18) to the low, intermediate and high risk subsets defined by unsupervised k-means clustering (Figure 5 and 6).

(B) Risk of any CT abnormalities, GGOs and lesions graded > five severity points at the 180 day follow-up (V3) in the risk subsets was assessed by unadjusted and acute COVID-19 severity-adjusted logistic regression models. Statistical significance of OR estimates was assessed by Wald Z test, p values were corrected for multiple comparisons by Benjamini-Hochberg method. Points with whiskers represent OR with 95% CI, dashed lines represent OR = 1. N = 108.

### Supplementary Figure S5

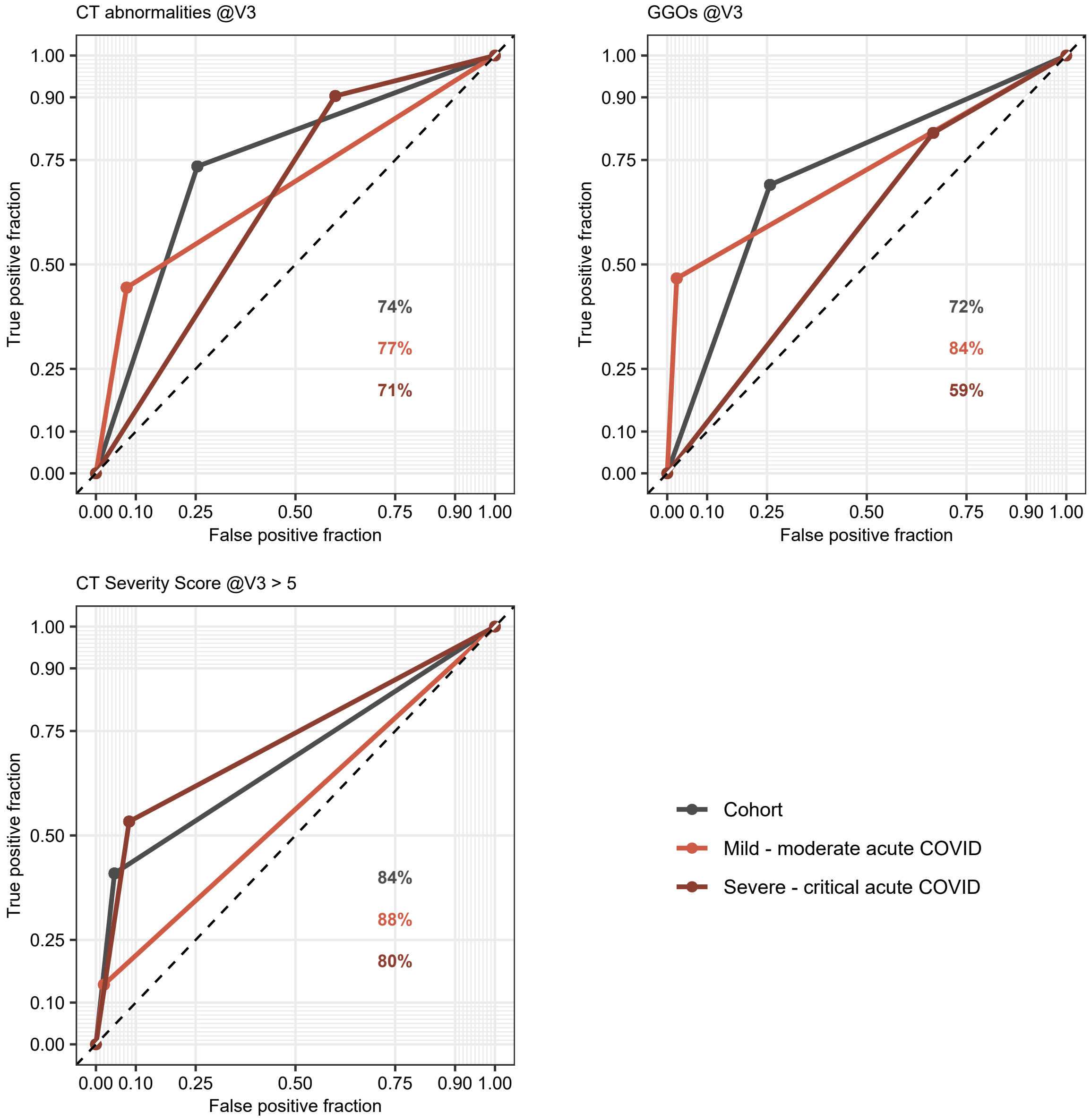


**Figure S5. Accuracy of prediction of radiological lung lesions at the 180 day follow-up by the kNN algorithm in in study participants stratified by acute COVID-19 severity.**

Occurrence of any CT lung abnormality, GGOs and CT lung lesions graded > five severity points at the 180 day follow-up (V3) was predicted by the weighted kNN algorithm for all study participants with a series of 108 one-out training/test cohort splits (training: n = 107, test = 1). The predicted outcome was compared with the true outcome in the entire cohort, patients with mild-to-moderate (outpatients and inpatients without ventilation, n = 57) and severe-to-critical (inpatients with oxygen therapy and ICU, n = 51) acute disease and visulaized as receiver-operator charcateristic (ROC) plots. Line color codes for the cohort subset, percents of correct preditions are indicated in the plots.

#### Supplementary Figure S6

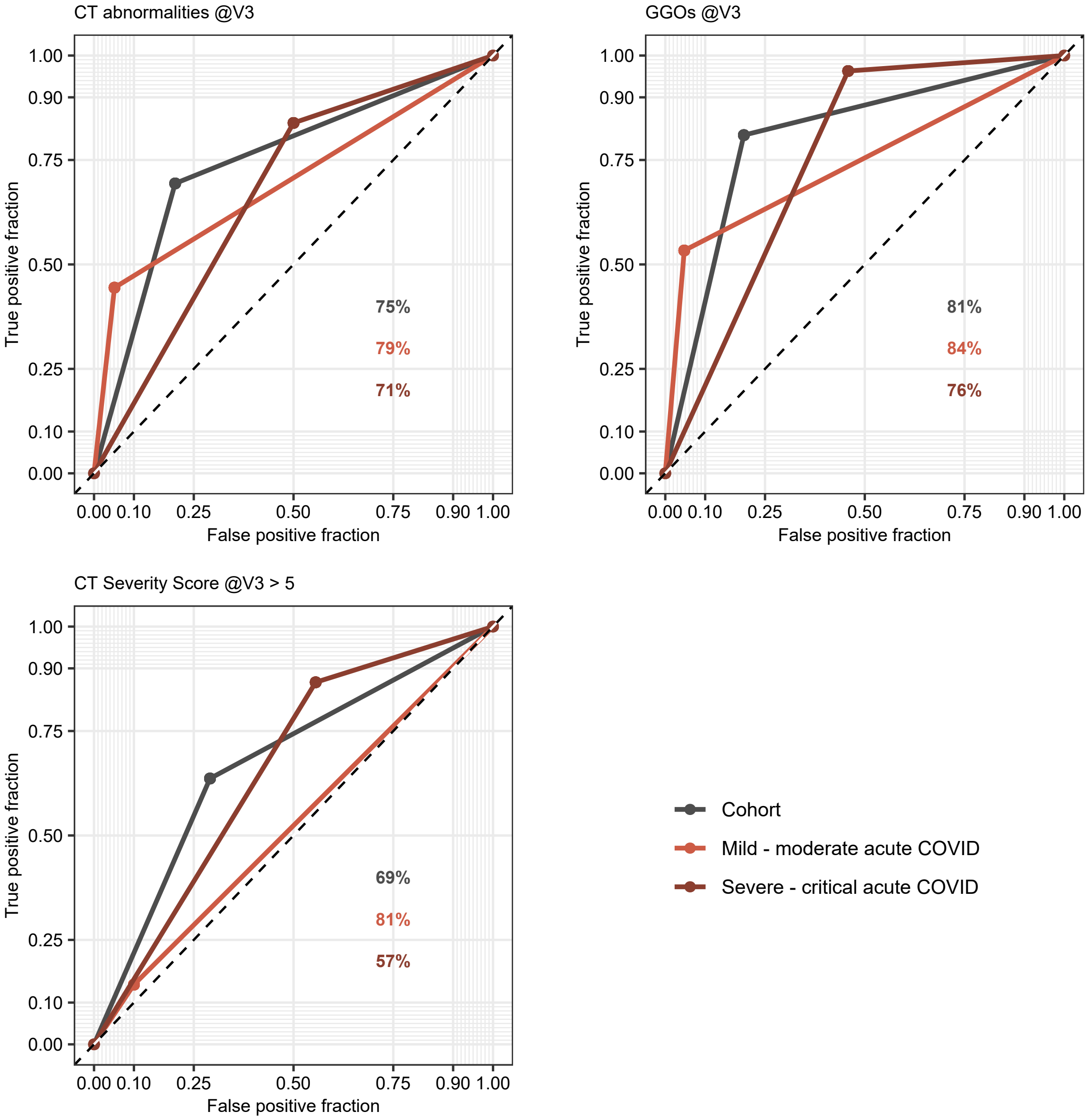


**Figure S6. Accuracy of prediction of radiological lung lesions at the 180 day follow-up by the naive Bayes algorithm in in study participants stratified by acute COVID-19 severity.**

Occurrence of any CT lung abnormality, GGOs and CT lung lesions graded > five severity points at the 180 day follow-up (V3) was predicted by the naive Bayes algorithm for all study participants with a series of 108 one-out training/test cohort splits (training: n = 107, test = 1). The predicted outcome was compared with the true outcome in the entire cohort, patients with mild-to-moderate (outpatients and inpatients without ventilation, n = 57) and severe-to-critical (inpatients with oxygen therapy and ICU, n = 51) acute disease and visulaized as receiver-operator charcateristic (ROC) plots. Line color codes for the cohort subset, percents of correct preditions are indicated in the plots.
